# Wastewater is Not a Reliable Proxy for Clinical Carbapenem Resistance Prevalence in *E. coli*

**DOI:** 10.64898/2026.10.02.26364587

**Authors:** Björn Berglund, Vaishnavi Divya Shridar, Marie Wrande, Jon Jerlström-Hultqvist, Stefan Engblom, Linus Sandegren, Sanna Koskiniemi

**Affiliations:** Department of Cell and Molecular Biology, Uppsala University, Sweden; Department of Information Technology, Uppsala University, Uppsala, Sweden; Department of Medical Biochemistry and Microbiology, Uppsala University, Uppsala, Sweden; Science for Life Laboratory, Department of Information Technology, Uppsala University, Uppsala. Sweden; Uppsala Antibiotic Center, Uppsala University, Uppsala, Sweden

## Abstract

Antimicrobial resistance (AMR) is a major threat to modern medicine and an emerging environmental pollutant. Wastewater-based monitoring offers a scalable alternative to traditional surveillance by enabling population-level monitoring across One Health compartments. However, it remains unclear how well bacterial variants detected in wastewater reflect those causing clinical infections. Here, we assessed the prevalence of carbapenem-resistant *Escherichia coli* (CREC) in wastewater from Uppsala, Sweden, and compared wastewater isolates with clinical isolates from the same region and period. We observed partial overlap between wastewater and clinical CREC populations, but several common clinical variants were not detected in wastewater and vice versa. CREC prevalence was an order of magnitude higher in wastewater receiving hospital effluent, yet this did not improve concordance with clinical isolates, suggesting that hospital wastewater may artificially inflate CREC prevalence estimates. These findings indicate that wastewater CREC prevalence does not necessarily reflect the regional clinical burden. Furthermore, metagenomic sequencing of wastewater-derived *E. coli* populations rarely detected CREC resistance genes, suggesting that CREC abundance is below the detection limit of this approach and that targeted enrichment is required for reliable monitoring. Together, our results show that while wastewater-based monitoring can complement AMR surveillance, it should be interpreted cautiously when used as an indicator of clinical risk.

## Introduction

Antimicrobial resistance (AMR) is a “silent pandemic” leading to increased mortality and healthcare costs worldwide. The number of deaths due to AMR in 2019 is estimated to 1.3–5.0 million [1], accounting for 2%–9% of all deaths during the year [2]. AMR monitoring is integral to provide actionable information, allowing preparedness against potential epidemic outbreaks and facilitating sustainable antibiotic usage [3]. However, healthcare-based surveillance describes the current clinical problem and cannot predict outbreaks driven by community-associated epidemiological shifts or risks arising from other sources, such as animal care or environmental transmission [4].

Wastewater-based monitoring (WBM) has emerged as a promising approach for surveillance of antimicrobial resistance (AMR). Because wastewater represents microorganisms from multiple One Health compartments [4, 5], WBM provides a cost-effective and less ethically challenging alternative to clinical or volunteer-based sampling and has already proven valuable for outbreak surveillance [6, 7]. Recognizing its potential, the European Commission has proposed EU-wide WBM to identify AMR transmission sources. However, the lack of standardized methodologies currently limits comparisons across regions and may compromise data quality. Establishing standardized protocols could make WBM a valuable tool for tracking population-level AMR and its clinical relevance.

A key unresolved question is how accurately AMR prevalence in wastewater reflects resistance patterns in the general population and clinical infections. While bacterial sequence types and AMR genes detected in wastewater often mirror those found in clinical settings [8-10], evidence linking identical isolates across both sources remains limited [11, 12]. Metagenomic studies have shown correlations between wastewater AMR gene abundance and resistance in clinical isolates, particularly bloodstream *Escherichia coli* isolates [13]. Hospital effluents also tend to contain a higher abundance and diversity of clinically relevant resistant bacteria than municipal wastewater [14], although further studies are needed to determine how frequently clinical and wastewater isolates overlap.

Carbapenems are last-resort antibiotics for treating infections caused by multidrug-resistant Gram-negative bacteria. Their effectiveness is increasingly threatened by the spread of plasmid-borne carbapenem resistance genes, and carbapenem-resistant *E. coli* (CREC) are now classified as critical priority pathogens by the World Health Organization [15, 16] [17]. Although studies of CREC in municipal wastewater remain limited, available data indicate that CREC are frequently detected in both high- and low-income settings [18]. In countries with high clinical prevalence, such as China, wastewater CREC have been shown to overlap with clinical isolates [19, 20]. However, it remains unclear whether wastewater prevalence reflects the clinical burden in low-endemicity settings.

Here, we investigated the prevalence and characteristics of CREC in wastewater from an urban region of Sweden, a low-endemicity country, over 12 months (2023-2024). Using selective cultivation and whole-genome sequencing, we characterized isolates by sequence type and antimicrobial resistance genes and compared their prevalence and clonality with local clinical isolates over time to evaluate the potential of wastewater-based monitoring for CREC surveillance in a low-endemicity setting.

## Materials and Methods

### Wastewater sample collection

Influent wastewater was collected monthly from July 2023 through June 2024 at a wastewater treatment plant in Uppsala, Sweden, which treats approximately 20 million m^3^ of wastewater annually and serves a population of approximately 333,000. Wastewater samples were collected as 48 h composite samples of 250 mL. Every month, one sample was collected from each of the two wastewater inlets AB and C. The wastewater samples were immediately stored at 4 °C and processed within 24 h of collection.

### Isolation and enumeration of carbapenem-resistant *Escherichia coli*

Wastewater was directly plated in duplicate on CHROMagar mSuperCARBA (CHROMagar, Saint-Denis, France) plates in 100 µL or 500 µL volumes. After incubation at 37 °C for 18-22 h, putative CREC colonies were morphologically identified according to the manufacturer’s instructions and enumerated. Each month, up to 10 CREC from each inlet (AB and C) were isolated and stored at -80 °C in 10% dimethyl sulfoxide (Sigma-Aldrich). For normalization of CREC counts, the number of total putative *E. coli* was enumerated after direct plating of tenfold serial dilutions (from 1:1 to 1:1000) of 100 µL wastewater on CHROMagar Orientation (CHROMagar), incubation at 37 °C for 18-22 h, and morphological identification according to the manufacturer’s instructions.

### Verification of carbapenem resistance

Verification of carbapenem resistance phenotypes was performed for 53 putative CREC isolated between July 2023 and June 2024 with antimicrobial susceptibility testing and minimal inhibitory concentration-(MIC)-determination by using E-tests (bioMérieux, Marcy-l’Étoile, France) for meropenem. In short, overnight cultures of the isolates grown in Luria-Bertani medium were adjusted to 0.5 McFarland and streaked on Mueller-Hinton agar (BD) plates. After application of the E-test-strip, the MICs were recorded after incubation at 37 °C for 18-22 h and evaluated according to the European Committee on Antimicrobial Susceptibility Testing (EUCAST) clinical breakpoints v 15.0.

### Genome sequencing and bioinformatic analyses

A total of 22 isolates were selected for whole-genome sequencing using nanopore. With the aim of sequencing one isolate from each inlet at each time-point, we selected isolates with MIC > 2mg/L Meropenem according to the EU-cast standard for susceptibility. In some instances, none of the isolates on the selective plates had a MIC that were high enough, in which case isolates with MIC between 0.25mg/L-1.5mg/L and were included in the analysis (Table S3).

DNA extraction was performed with the Genomic-tip 100/G (QIAGEN, Hilden, Germany) kit according to the manufacturer’s instructions. Quality and quantity of the extracted DNA was evaluated by using a Qubit 2.0 Fluorometer (Invitrogen, Waltham, MA) with the Qubit dsDNA HS assay kit (Thermo Fisher Scientific) and a NanoDrop Spectrophotometer (Thermo Fisher Scientific). Genome libraries were prepared by using the Rapid sequencing DNA V14 – barcoding kit (Oxford Nanopore Technologies, Oxford, UK) according to the manufacturer’s instructions. Long-read sequencing was performed on an R10.4 flow cell by using MinION Mk1C running MinKNOW v24.11.8 software. The raw Nanopore data were basecalled using Dorado v0.9.1 in stereo duplex super-accuracy mode using the dna_r10.4.1_e8.2_400bps_sup@v5.0.0 and dna_r10.4.1_e8.2_5khz_stereo@v1.3 models. Basecalled reads were binned by dorado demux using kit SQK-RBK-114-96. Adapters were trimmed using dorado trim and converted to fastq format using samtools fastq. One additional round of adapter and chimera removal was done by using Porechop v0.2.3 [21]. The Nanopore reads were filtered by using Filtlong v0.2.1 [22] to discard reads less than 1000 bp and 10% of reads of the lowest quality. The Nanopore datasets were assembled by using Flye v2.9.2-b1786 [23] with the --nano-hq flag, two rounds of polishing, --no-alt-contigs and the completed assemblies were inspected by Bandage [24]. The assemblies were polished by dorado polish. To this end, reads rebasecalled in simplex mode with --emit-moves, trimmed using dorado trim and aligned using dorado aligner. The resulting bam-file was used to polish the assemblies using dorado polish with the --bacteria flag The assemblies were annotated by bakta webserver v.1.11.0 using the v6.0 full database [25, 26].

A total of 22 isolates, representing all isolates found on the same day from the same inlet on three different occasions during the sampling period (230828, 240130, 240626) were selected for Illumina sequencing. Genomic DNA was prepared from 22 isolated colonies using the MasterPure complete DNA and RNA purification kit (Nordic Biolabs, Sweden). PCR-free library construction and whole-genome re-sequencing was performed by the BGI sequencing facility (Hong Kong, China) using the HiSeq 4000 PE150 sequencing system (Illumina Inc., San Diego, CA, USA). Illumina analysis pipeline was used for image analysis, base calling and quality score calibration. Raw sequence reads were filtered and exported as FASTQ files, which were assembled into contigs using the built-in pipeline in the CLC genomics workbench (CLC, Denmark).

AMR genes were evaluated using AMRFinderPlus v4.0.23 with the NCBI Reference Gene Database v2025-07-16.1. The presence of *bla*_NDM-5_, *bla*_OXA-181_ and *bla*_OXA-244_ was specifically assessed using BLASTN v2.16.0+ [27].

### Clinical CREC isolates

To compare the whole-genome sequenced isolates above against CREC reported by the Public Health Agency of Sweden between 2023 and 2024, genomes from CREC isolated from144 patient from clinics in the Uppsala/Stockholm region 1 with labelled Achtman ST and ESBL genes were obtained. Raw reads were quality- and length-filtered using fastp v1.0.1 (minimum Phred score = 20, minimum length = 50 bp), with adaptive trimming applied at both read ends using a sliding window approach (window size = 4, mean quality threshold = 20) [28]. Contigs were assembled from the trimmed reads using SPAdes v4.0.2 with the --assembler-only and --careful flags to reduce the number of mismatches and short indels [29].

To evaluate the relatedness between these isolates, autoMLST2.0 was used for gene selection, core gene alignment, and outgroup identification with the flags -wf 1 -mo 0 -cat -c 12 [30]. Phylogenetic inference was performed using IQ-TREE (-st DNA, -m GTR+F, -bb 1000, -alrt 1000), employing maximum likelihood with model selection and branch support estimated via ultrafast bootstrap and SH-aLRT [31]. The resulting phylogenetic tree was drawn in R v4.5.1 using ggtree [32]. Genome similarity across eight clusters identified from the phylogenetic tree was assessed using pyANI-plus v1.0.0 [33]. This analysis was repeated for the Illumina sequenced genomes from the same inlet on the same day.

### Metagenomic sequencing and analysis of metagenomic data

Based on a mixture of ∼1000 colonies from the *E.coli* selective plates, DNA was prepared and sent for metagenomic sequencing. Raw sequencing reads were first processed with fastp v1.0.1 to remove adapter sequences and perform quality filtering [28]. Host-derived reads were subsequently removed by alignment to the human reference genome using Bowtie2 v2.5.2 [34]. The resulting non-host reads were taxonomically profiled using MetaPhlAn v3.1.0 with the mpa_v31_CHOCOPhlAn_201901 database to estimate microbial taxon relative abundances [35]. The presence of *bla*_NDM-5_, *bla*_OXA-181_ and *bla*_OXA-244_ in the produced raw metagenomes collected in the same time period was assessed using bwa-mem2 v2.3 [36].

## Results

### Prevalence of carbapenem-resistant *Escherichia coli* in wastewater from Uppsala

Throughout the one-year sampling period (July 2023 to June 2024), colonies of putative *E. coli* and CREC were consistently cultivable from wastewater samples collected at both inlets AB and C at the Uppsala wastewater treatment plant (Fig. 1AB). Inlet AB combines wastewater from both the Uppsala community as well as Uppsala University Hospital, whereas inlet C only contains wastewater from the community (Fig. 1A). Only two samples, collected in September and December 2023, respectively (both from inlet C) were negative for growth of putative CREC (Fig. 1C). This is likely because the numbers of CREC were close to the detection limit (hatched line Figure 1C) for the C inlet throughout the time period. The mean number of total putative *E. coli* was 8.2×10^6^ cfu / 100 mL (SEM: 1.2×10^6^) in inlet AB and 1.0×10^7^ cfu / 100 mL (SEM: 1.5×10^6^) in inlet C, respectively (Fig. 1C, Table S1). The mean number of CREC was 1.5×10^4^ cfu / 100 mL (SEM: 4×10^3^) in inlet AB and 3.1×10^3^ cfu / 100 mL (SEM: 1.1×10^3^) respectively (Fig. 1C, Table S1). Thus, the prevalence of CREC in relation to total *E.coli* was rather stable throughout the sampling period 0.34+/- 0.21% CREC of total *E. coli* for inlet AB and 0.02+/-0.01% for inlet C. The prevalence of putative CREC was 5 times higher in the inlet containing hospital wastewater (P=0.0133).

**Figure 1.**
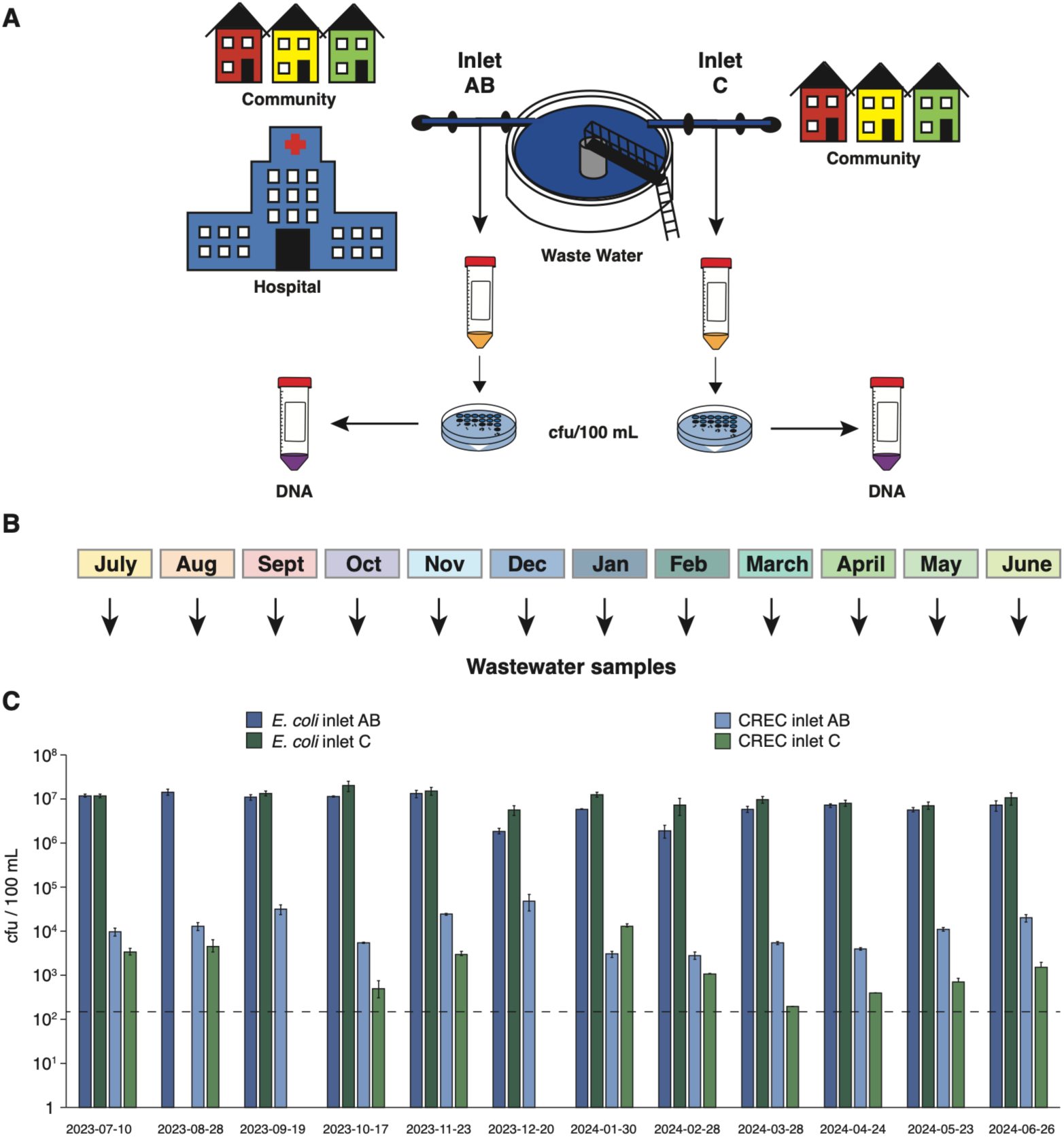
Prevalence of total *Escherichia coli* and carbapenem-resistant *E. coli* in wastewater from Uppsala, Sweden. A) Overview of sampling set-up. B) Sampling period. C) The prevalence of total *E. coli* (dark colours) and carbapenem-resistant *E. coli* (CREC) (lighter colors) in Uppsala wastewater from two inlets AB (blue) and C (green). Error-bars represent SEM.

### Genetic diversity of CREC from wastewater

A total of 21 CREC isolates were selected for whole-genome long-read sequencing with the Oxford Nanopore platform. These isolates represented 8 different sequence types (STs), where ST361 was the most common (n=7), followed by ST167 (n=4), ST410 (n=3), and ST90 and ST648 (n=2). One isolate each was found to belong to ST38, ST1284, and ST1300, respectively whereas one isolate could not be verified due to low read quality (Fig. 2, Table S1). All isolates were found to carry either of carbapenem resistance genes *bla*_NDM-5_ (n=13), *bla*_OXA-181_ (n=5), *bla*_OXA-244_ (n=1) or a combination of *bla*_NDM-5_ and *bla*_OXA-181_ (n=3) (Fig. 2, Table S2, S3). In addition, the isolates carried a diverse and varied range of AMR genes for other antibiotics including aminoglycosides, cephalosporins, phenicols, macrolides, quinolones, sulphonamides, trimethoprims, and tetracyclines, with *aadA2* (n=15), *mph(A)* (n=18), *sul1* (n=19), *dfrA12* (n=14), and *tet(A)* (n=13) being the most common (Table S2). 8 isolates carried the ESBL-gene *bla*_CTX-M-15_, and 10 isolates carried *bla*_CMY-42_ (n=6) or *bla*_CMY-145_ (n=4) (Table S2).

**Figure 2.**
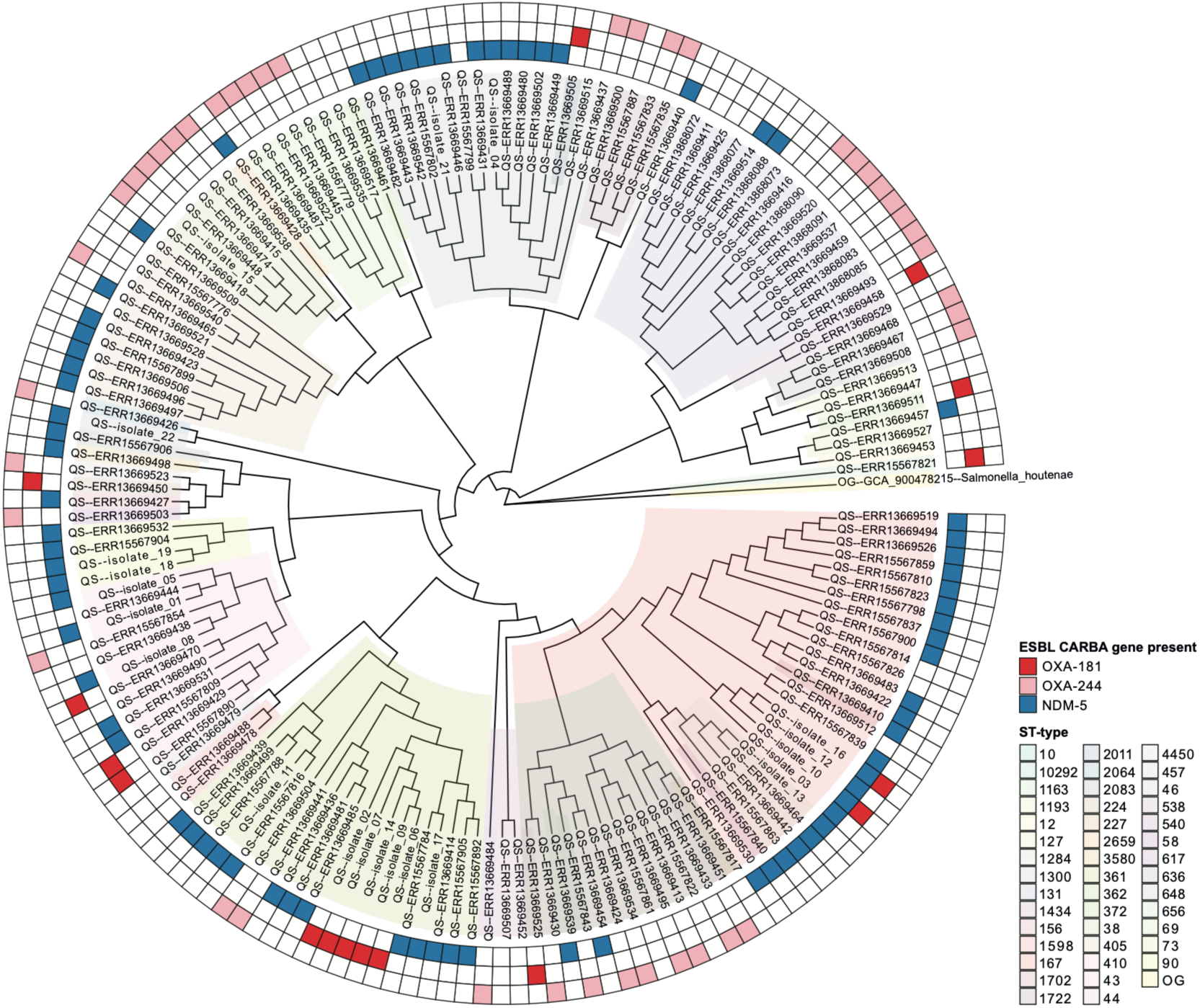
Carbapenem-resistant *Escherichia coli* (CREC) from the clinic and wastewater in the Uppsala-Stocholm region, Sweden. Phylogenetic tree illustrating the CREC isolated in the clinic from Uppsala-Stocholm between 2023-2024 and whole-genome sequenced CREC isolated in this study isolated between July 2023 and August 2024 from the Uppsala region. Antibiotic resistance genes identified in the isolates are shown as boxes outside of the variant in the tree. ST-types are indicated by colour according to the legend on the right.

### Poor correlation between CREC isolated from wastewater and clinic

To assess whether wastewater can be used for prediction or real-time monitoring of AMR disease burden, it is important to determine whether wastewater isolates are clonally related to those circulating in the clinic. Because resistance genes may be carried by commensal bacteria that do not cause infection, their presence alone may not reflect the clinical burden. Therefore, genomes of all clinical CREC isolates from the Uppsala/Stockholm region (n = 144) were downloaded from the Public Health Agency of Sweden. Inclusion of the wider region was motivated by extensive commuting (>40,000 persons/day), whereby individuals contributing to wastewater in Uppsala may seek healthcare in Stockholm. Since clinical detection may occur before or after detection in wastewater, clinical isolates collected through December 2024 were included (Fig. S1).

To assess relatedness, we constructed a phylogenetic tree of clinical and wastewater isolates using *Salmonella houtenae* (GCA_900478215) as an outgroup. The tree grouped isolates into at least 15 distinct clusters with little or no phylogenetic distance within clusters (Fig. S2). Eight clusters contained both clinical and wastewater isolates, suggesting close relatedness between the two sources (Fig. S2). Nineteen of 22 sequenced wastewater isolates belonged to clusters that also contained clinical isolates from the study period (Fig. S2, Table S4). Two wastewater isolates collected from the same outlet at the same time were identical and were therefore treated as a single isolate in subsequent analyses.

Pairwise genome comparisons within clusters showed that 12 of 21 unique wastewater CREC isolates matched clinical isolates using a threshold of <2,000 SNPs, while a further six matched using a threshold of <5,000 SNPs (Table S4). Three matching isolates were detected in the clinic both before and after detection in wastewater, whereas six and nine were detected only before or only after wastewater detection, respectively. In these cases, closely related isolates (<10,000 SNPs) were present both before and after wastewater detection (Table S4). Matching isolates originated from both inlets (56% from AB and 44% from C) (Table S4). Isolates from inlet AB were more often detected in the clinic after wastewater detection (88%) than isolates from inlet C (29%). Together, these findings indicate that CREC variants circulating in the clinic can often be detected in wastewater during the same period, although some variants were found only in wastewater or only in clinical samples.

### Genes encoding carbapenem resistance are not detectable in metagenomes from wastewater or *E. coli* enriched populations

The European Commission has proposed wastewater-based monitoring (WBM) as a tool for AMR pandemic preparedness [37]. Its utility depends on whether clinically relevant AMR variants can be reliably detected in wastewater and occur at sufficient prevalence. One strategy to improve detection may be to enrich for key pathogens such as *E. coli*. In this low-endemicity region, CREC represented only 0.3% of the *E. coli* population when hospital wastewater was included and 0.04% in community wastewater, raising the question of whether carbapenem resistance genes can be detected by metagenomic sequencing of wastewater or *E. coli*-enriched samples.

To address this, DNA was extracted from wastewater samples that yielded sequenced CREC isolates, as well as from *E. coli*-enriched populations generated by pooling approximately 1,000 putative *E. coli* colonies from each sample (24 samples; two inlets sampled at 12 time points). Metagenomic sequencing was performed on DNA from the original wastewater and the enriched populations. Wastewater DNA from three time points and *E. coli*-enriched DNA from 12 time points with detected CREC were sequenced. Metagenomes were screened for the carbapenem resistance genes identified in the wastewater isolates, *bla*_NDM-5_, *bla*_OXA-244_, and *bla*_OXA-181_. Among the 21 isolates, 25 carbapenem resistance genes were detected, with most isolates carrying a single gene and three carrying two genes.

None of the target genes were detected in the wastewater metagenomes (Table S2). In the *E. coli*-enriched metagenomes, where *E. coli* accounted for 75-97% of the reads (Table S2), no reads mapping to the corresponding resistance genes were detected in 16 of 22 samples (Fig. 3). Among samples containing isolates with multiple resistance genes, one gene was detected in two metagenomes, whereas no genes were detected in the third (Fig. 3, Table S2). Collectively, these findings indicate that reliable estimation of CREC prevalence in this low-endemicity setting is not feasible using wastewater or *E. coli*-enriched metagenomic sequencing alone.

**Figure 3.**
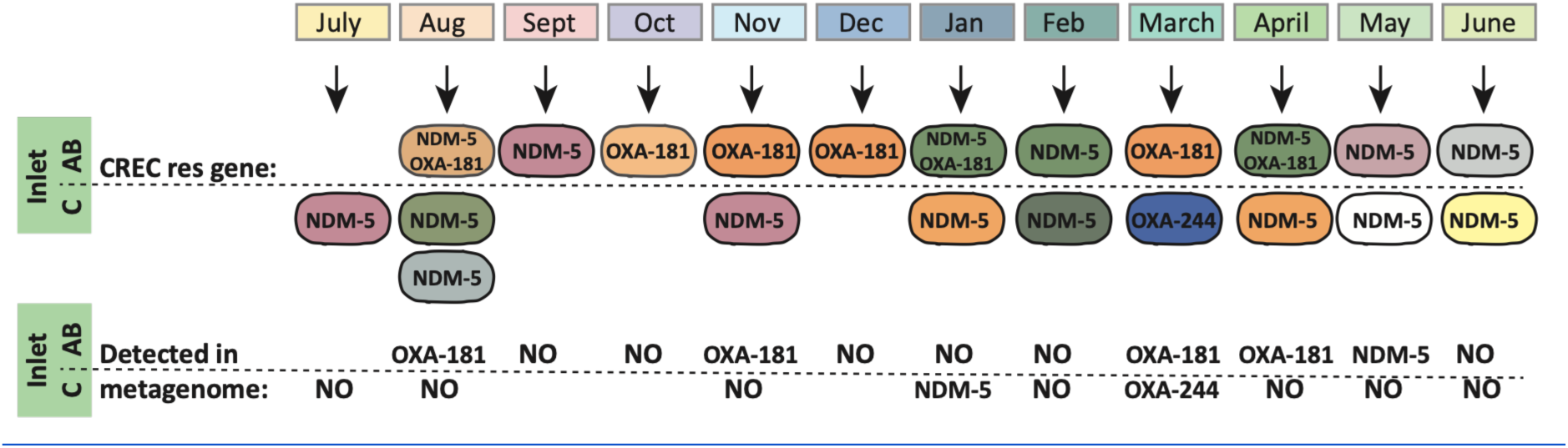
Isolation of carbapenem-resistant *Escherichia coli* (CREC) from wastewater. Overview of isolates selected for whole-genome sequencing from the sampling period. Similar colours indicate genetic similarity. Identical colours indicate <2000SNPs difference. The carbapenem resistance genes (e.g. *bla*_NDM-5_ (NDM-5) or *bla*_OXA-181_(OXA-181)) found in these isolates are shown in the table below the illustration. Detection of carbapenem resistance genes in the *E. coli* enriched metagenomes is indicated below each sample.

## Discussion

Here, we show that wastewater reflects community carriage of clinical CREC isolates reasonably well at the isolate level, but that inclusion of hospital effluent can inflate the apparent prevalence of resistant bacteria. Only 57% of sequenced wastewater CREC isolates matched sequence types detected in clinics from the same region and period. Notably, these isolates originated from both wastewater inlets and not only the inlet receiving hospital waste. Because hospital effluents are often enriched in clinically relevant resistant bacteria [38], the higher CREC prevalence observed in the hospital-associated inlet suggests that many CREC isolates originated from the hospital. As most clinical isolates from the region are also collected at the hospital, a stronger correlation between clinical and wastewater isolates would be expected in this inlet.

One explanation is that carbapenem use, which is largely restricted to hospitals, selects for CREC within patients or the hospital wastewater environment. Alternatively, carbapenem treatment of patients already colonized with CREC may increase fecal shedding and consequently CREC concentrations in wastewater. Most isolates collected from the same inlet on the same day were clonal (Table S3), supporting selection at some level. Supporting selection in patients specifically, 88% of matching isolates were detected in hospital containing wastewater before clinical isolation. Thus, these CREC isolates became detectable in the clinic after wastewater detection, suggesting enrichment in the hospital and not in the water. Together, these observations are more consistent with enrichment in colonized patients than selection within the wastewater itself. If so, the elevated CREC prevalence in hospital-associated wastewater may not reflect a higher prevalence in the hospital population than in the surrounding community. Further work is therefore needed to determine the source of the increased CREC prevalence in wastewater containing hospital effluent. Clarifying this issue could inform infection control and antimicrobial stewardship strategies, including recommendations for hospital wastewater treatment or rapid AMR profiling before antibiotic administration. Until then, hospital effluent should be excluded from wastewater-based community surveillance, as its inclusion may overestimate CREC prevalence in the general population.

Most CREC variants detected in wastewater could also be identified among clinical isolates before and after their detection in wastewater when a relatedness threshold of <10,000 SNPs was applied, indicating substantial overlap between clinical and wastewater populations. Despite this, clinically relevant variants occurred at frequencies too low for reliable detection in unenriched metagenomic datasets. Although the number of wastewater CREC isolates was limited, three isolates had no corresponding clinical match, suggesting the presence of a community reservoir not captured by clinical surveillance. Conversely, several common clinical variants were never detected in wastewater. Thirteen clusters of closely related clinical isolates were absent from all wastewater samples, despite seven of these being repeatedly isolated over periods exceeding five months. This was comparable to the clusters detected in wastewater, which were repeatedly recovered over >5 months in seven of eight cases. The discrepancy therefore cannot be explained solely by transient circulation and may instead reflect differences in survival, persistence, or recovery in the wastewater environment between isolates. Such biases could potentially be reduced by culture-independent approaches that do not rely on recovery of viable bacteria, and many AMR surveillance efforts are moving in this direction. However, our results suggest that in low-endemicity settings such as Sweden, clinically important variants carrying resistance to last-resort antibiotics occur below the detection limit of wastewater metagenomics. As a result, culture-independent approaches may fail for the same reason as culture-based methods: the target organisms are too rare. Collectively, these findings question the utility of wastewater surveillance as a standalone approach for monitoring clinically relevant CREC and other low-prevalence AMR pathogens.

## Data Availability Statement

All raw data for the manuscript are freely available either as supplementary excel files (Tables S1 and 4) or at public repositories. The whole genome sequenced isolates can be found at https://figshare.com/s/a940d2b8aa0ccf405810 whereas the metagenomes have been deposited at https://www.ebi.ac.uk/biostudies/studies/S-BSST3367.

## Author Contributions

B.B, V.D.S., M.W., S.E., L.S., and S. K. conceived the study. B.B, V.D.S., M.W. and J.J.H. performed experiments. B.B, V.D.S., M.W., J.J.H., S.E., L.S., and S.K analyzed data. B.B, V.D.S., S.E., L.S., and S. K wrote the manuscript.

## Acknowledgements

We acknowledge the National Academic Infrastructure for Supercomputing in Sweden (NAISS), partially funded by the Swedish Research Council through grant agreement no. 2022-06725, for awarding this project access to NSC Tetralith for the performing of computations.

## Ethical considerations

Ethical review and informed consent were not required for this study because all wastewater samples were collected at the population level and could not be linked to identifiable individuals. No personal or clinical information was collected or accessed. Clinical bacterial genomes included in the analyses were obtained from publicly available repositories and contained no patient-identifiable information. The study does not include randomised controlled trials nor economic evaluations of health interventions.

