## Supplementary files for "Wastewater is Not a Reliable Proxy for Clinical Carbapenem Resistance Prevalence in *E. coli*"

**This file contains:**

**Supplementary figures 1-2**

**Supplementary tables 1-4**

**Figure S1. Overview of timeline.** Timeline of sampling from wastewater and clinic is shown. Wastewater sampling occurred between July 2023 and June 2024 (12 months). The clinical isolates included were from AB and C regions and collected between January 2023 and December 2024.

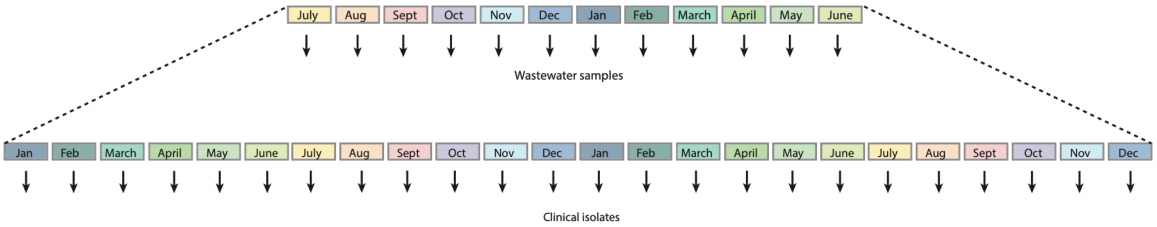

**Figure S2. Phylogenetic tree of isolates from clinic and wastewater.** Phylogenetic tree of carbapenem resistant E.coli isolates collected in the clinic during January 2023 – 2024 December and wastewater isolates from July 2023 – 2024 June. Isolates of the same evolutionary distance to outgroup are marked with pink color and referred to as clusters 1-X.

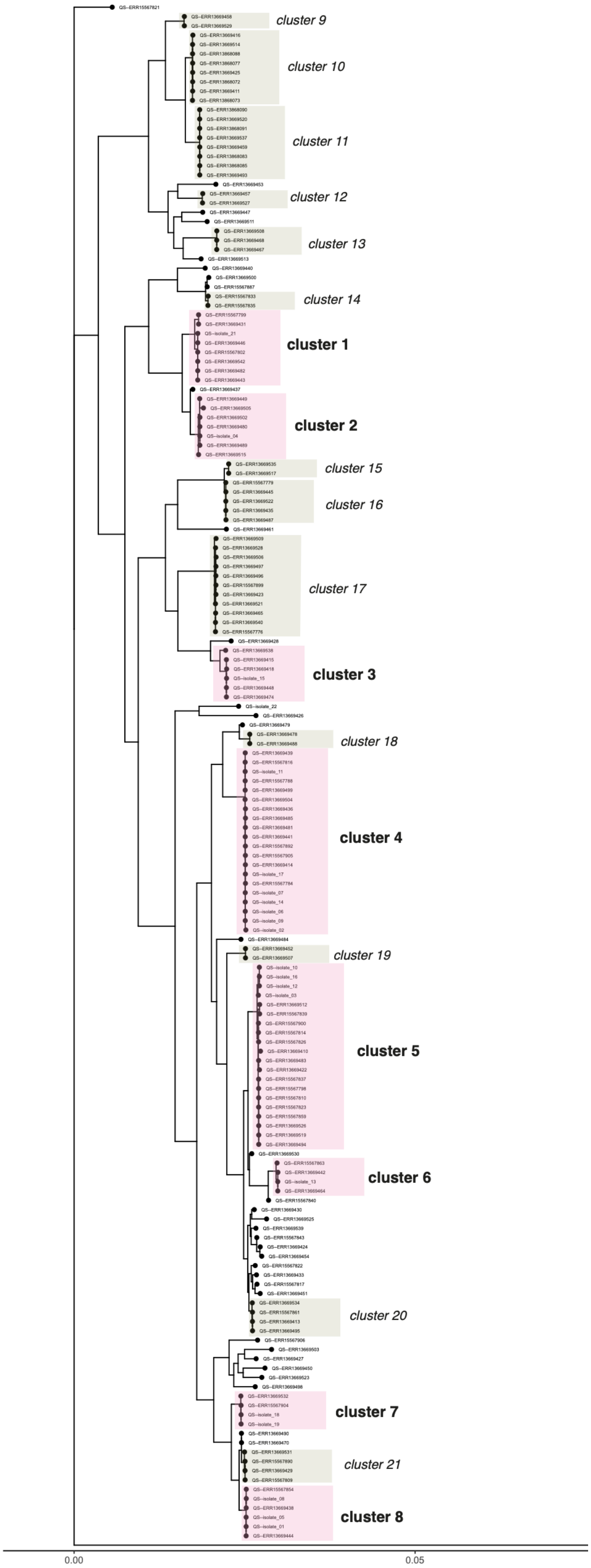

**Table S1. Raw data and bioinformatic analyses of the sequenced genomes.**

Table S1 is provided as a separate excel file.

**Table S2.** Antibiotic resistance genes and plasmid distribution of 14 carbapenem-resistant *Escherichia coli* isolated from urban wastewater in Sweden. Carbapenem resistance genes are indicated in bold.

| Isolate | Name | Resistance genes |
|---|---|---|
| **Isolate 1** | **CEC4** | *aadA2*, ***bla*NDM-5**, *mph(A)*, *sul1*, *dfrA12, bla*CMY-42, *aadA2*, *blaTEM-1B*, ***bla*NDM-5**, *mph(A)*, *sul1*, *tet(A)*, *dfrA12* |
| **Isolate 2** | **AB2309-1** | *aadA1*, *blaOXA-1*, *catA1*, *qepA1*, *aadA5*, *mph(A)*, *sul1*, *tet(B)*, *dfrA17*, ***blaOXA-181*** |
| **Isolate 3** | **C2309-1** | *aadA2*, ***bla*NDM-5**, *mph(A)*, *sul1*, *dfrA12, bla*CMY-42 |
| **Isolate 4** | **C2309-4** | *aadA2*, ***bla*NDM-5**, *sul1*, *dfrA12* |
| **Isolate 5** | **AB23Sep-5** | *bla*CMY-42, *aadA2*, *bla*TEM-1B, ***bla*NDM-5**, *mph(A)*, *sul1*, *tet(A)*, *dfrA12* |
| **Isolate 6** | **A1023-4** | *ant(3")-Ia, blaOXA-1, blaOXA-1, qepA1, catA1, sul1, aadA5, dfrA17, mph(A), tet(B),* ***blaOXA-181*** |
| **Isolate 7** | **A1123-2** | *catA1, qepA1, blaOXA-1, ant(3")-Ia, qnrS1,* ***blaOXA-181****, balTEM-1C, catA2, tet(B), mph(A), dfrA17, aadA5, sul1* |
| **Isolate 8** | **C1123-1** | *aadA2, aac(6')-Ib-cr, blaCTX-M-15, blaOXA-1,* ***blaNDM-5****, mph(A), catB3, sul1, tet(A), dfrA12, blaCMY-42, blaTEM-1B* |
| **Isolate 9** | **A1223-6** | *aadA1, blaOXA-1, catA1, qepA1,* ***blaOXA-181****, qnrS1, catA2* |
| **Isolate 10** | **A0124-3** | *blaCMY-145, qnrS1****, blaOXA-181****,* ***blaNDM-5****, sul1, aadA2, dfrA12, blaCTX-M-15, blaOXA-1, aac(6")-Ib-cr, tet(A)* |
| **Isolate 11** | **C0124-1** | *mph(A),* ***blaNDM-5,*** *sul1, aadA2, dfrA12, tet(A), blaCTX-M-15* |
| **Isolate 12** | **A0224-2** | *aadA2*, *aac(6')*-Ib-cr, *bla*CTX-M-15, *blaOXA-1*, ***bla*NDM-5**, *mph(A)*, *catB3*, *sul1*, *tet(A)*, *dfrA12*, *bla*CMY-145 |
| **Isolate 13** | **C0224-3** | *aac(3)*-IIa, *aac(6')*-Ib-cr, *aadA5*, *bla*CTX-M-15, *blaOXA-1*, ***bla*NDM-5**, *mph(A)*, *catB3*, *sul1*, *tet(B)*, *dfrA17* |
| **Isolate14** | **A0324-1** | *aadA1*, *blaOXA-1*, *catA1*, *qepA1*, *aadA5*, *mph(A)*, *sul1*, *tet(B)*, *dfrA17*, ***blaOXA-181***, *qnrS1* |
| **Isolate 15** | **C0324-1** | ***blaOXA-244,*** *dfrA17, aadA5, sul1, mph(A), sul2, aph(3")-Ib, aph(6)-Id, tet(A),* ***blaCTX-M-27*** |
| **Isolate 16** | **A0424-4** | *aadA2*, *aac(6')*-Ib-cr, *bla*CTX-M-15, *blaOXA-1*, ***bla*NDM-5**, *mph(A)*, *catB3*, *sul1*, *tet(A)*, *dfrA12*, ***blaOXA-181***, *qnrS1*, *bla*CMY-145 |
| **Isolate 17** | **C0424-1** | *blaCMY-145, ant(3")-Ia, blaOXA, qepA1, catA1,* ***blaNDM-5****, sul1, aadA2, dfrA12, mph(A)* |
| **Isolate 18** | **A0524-1** | *aadA2*, ***bla*NDM-5**, *mph(A)*, *sul1*, *tet(A)*, *dfrA12*, *bla*CMY-42 |
| **Isolate 19** | **A0524-6** | *aadA2*, ***bla*NDM-5**, *mph(A)*, *sul1*, *tet(A)*, *dfrA12*, *bla*CMY-42 |
| **Isolate 20** | **C0524-1_1** | *aadA2, aadA1,* ***bla*NDM-5**, *bla*OXA-1, *bla*CTX-M-15*, aac(6')-Ib-cr, mph(A), catB3, sul1, tet(A), dfrA12* |
| **Isolate 21** | **A0624-1** | *blaTEM-1B, aac(3)-Iia, blaOXA, aac(6")-Ib-cr, blaCTX-M-15, mph(A), tet(A), dfrA12, aadA2, sul1****, blaNDM-5****, sul1, aadA2, dfr12* |
| **Isolate 22** | **C0624-6** | *aadA2*, *bla*TEM-1b, ***bla*NDM-5**, *mph(A)*, *qepA4*, *sul1*, *tet(B)*, *dfrA12*, *bla*TEM-146, *floR*, *qnrS1*, *tet(A)*, *aph(6)*-Id, *aph(3")*-Ib |
| | **AB2309-2_1** | *aadA2,* ***bla*NDM-5**, *mph(A), bla*CMY-145, *sul1, tet(A), dfrA12* |
| | **AB2309-3_1** | *aadA5, bla*TEM-1B, ***bla*OXA-181**, *bla*OXA-1, *ant(3")-Ia, catA1, qepA1, sul1, tet(B), dfrA17* |
| | **AB2309-4_1** | *aadA5, bla*TEM-1C, *bla*OXA-1, ***bla*OXA-181**, *mph(A), ant(3")-Ia, catA1, qnrS1, qepA1, tet(B), dfrA17* |
| | **AB2309-5_1** | *aadA2, bla*CMY-42, ***bla*NDM-5**, *mph(A), sul1, tet(A), dfrA12* |
| | **AB2309-6_1** | *aadA5,* ***bla*OXA-181**, *bla*OXA-1, *mph(A), ant(3")-Ia, catA1, qepA1, sul1, tet(B), dfrA17* |

| | | |
|---|---|---|
| | **AB2309-7_1** | *aadA5, bla*TEM-1C*, bla*OXA-1*,* ***bla*OXA-181***, mph(A), ant(3'')-Ia, catA1, qepA1, qnrS1, tet(B), dfrA17* |
| | **A0124-1_1** | *aadA5, bla*TEM-1C*,* ***bla*OXA-181***, mph(A), qnrS1, sul1, tet(B), dfrA17* |
| | **A0124-4_1** | *aadA5,* ***bla*OXA-181***, bla*TEM-1B*, bla*OXA-1*, ant(3'')-Ia, catA1, qepA1, sul1, tet(B), dfrA17* |
| | **A0124-5_1** | *aadA5,* ***bla*OXA-181***, bla*TEM-1B*, bla*OXA-1*, ant(3'')-Ia, catA1, qepA1, qnrS1, sul1, tet(B), dfrA17* |
| | **A0124-7_1** | *aadA5, bla*OXA-1*, bla*TEM-1B*,* ***bla*OXA-181***, ant(3'')-Ia, catA1, qepA1, sul1, tet(B), dfrA17* |
| | **A0124-8_1** | *aadA5, bla*TEM-1B*, bla*OXA-1*,* ***bla*OXA-181***, ant(3'')-Ia, catA1, qepA1, qnrS1, sul1, tet(B), dfrA17* |
| | **A0124-9_1** | *aadA5, bla*OXA-1*,* ***bla*OXA-181***, mph(A), ant(3'')-Ia, catA1, sul1, tet(B), dfrA17* |
| | **A0124-10_1** | *aadA5, bla*TEM-1B*, bla*OXA-1*,* ***bla*OXA-181***, ant(3'')-Ia, catA1, qepA1, sul1, tet(B), dfrA17* |
| | **A0624-1_1** | *aadA2, aac(3)-IIa, bla*OXA-1*,* ***bla*NDM-5***, bla*CTX-M-15*, bla*TEM-1B*, aac(6')-Ib-cr, mph(A), catB3, sul1, tet(A), dfrA12* |
| | **A0624-2_1** | *aadA5, bla*OXA-1*,* ***bla*OXA-181***, bla*TEM-1B*, ant(3'')-Ia, catA1, qnrS1, qepA1, tet(B), dfrA17* |
| | **A0624-3_1** | *aadA5, bla*TEM-1C*, bla*OXA-1*,* ***bla*OXA-181***, mph(A), ant(3'')-Ia, catA1, qepA1, sul1, tet(B), dfrA17* |
| | **A0624-4_1** | ***bla*OXA-244** |
| | **A0624-5_1** | *aadA5,* ***bla*OXA-181***, bla*TEM-1B*, bla*OXA-1*,* ***bla*OXA-500***, ant(3'')-Ia, catA1, qnrS1, qepA1, tet(B), dfrA17* |
| | **A0624-6_1** | *aadA5, bla*OXA-1*, bla*TEM-1B*,* ***bla*OXA-181***, ant(3'')-Ia, catA1, qepA1, qnrS1, tet(B), dfrA17* |
| | **A0624-7_1** | *aadA5, bla*OXA-1*,* ***bla*OXA-181***, mph(A), ant(3'')-Ia, catA1, qepA1, qnrS1, sul1, tet(B), dfrA17* |
| | **A0624-8_1** | *bla*CMY-150*,* ***bla*OXA-181***, aph(3'')-Ib, aph(6)-Id, blaLAP-2, floR, qnrS1, qnrB38, sul2, dfrA14* |
| | **A0624-9_1** | *aadA5, bla*OXA-1*, bla*TEM-1B*,* ***bla*OXA-181***, ant(3'')-Ia, catA1, qepA1, qnrS1, sul1, tet(B), dfrA17* |

**Table S3. MIC of meropenem for sequenced wastewater isolates:**

| Isolate | Carbapenem resistance gene | MIC (meropenem) mg/L |
|---|---|---|
| Isolate 1 | *bla*$_{NDM-5}$ | 32 |
| Isolate 2 | *bla*$_{OXA-181}$, *bla*$_{NDM-5}$ | 1.5 |
| Isolate 3 | *bla*$_{NDM-5}$ | >32 |
| Isolate 4 | *bla*$_{NDM-5}$ | 6/8/12 |
| Isolate 5 | *bla*$_{NDM-5}$ | 24 |
| Isolate 6 | *bla*$_{OXA-181}$ | 1.5 |
| Isolate 7 | *bla*$_{OXA-181}$ | 1.5 |
| Isolate 8 | *bla*$_{NDM-5}$ | >32 |
| Isolate 9 | *bla*$_{OXA-181}$ | 0.25 |
| Isolate 10 | *bla*$_{NDM-5}$, *bla*$_{OXA-181}$ | 8 |
| Isolate 11 | *bla*$_{NDM-5}$ | 10 |
| Isolate 12 | *bla*$_{NDM-5}$ | 4 |
| Isolate 13 | *bla*$_{NDM-5}$ | 0.75 |
| Isolate 14 | *bla*$_{OXA-181}$ | 0.25 |
| Isolate 15 | *bla*$_{OXA-244}$ | 0.25 |
| Isolate 16 | *bla*$_{OXA-181}$, *bla*$_{NDM-5}$ | 16 |
| Isolate 17 | *bla*$_{NDM-5}$ | 24 |
| Isolate 18 | *bla*$_{NDM-5}$ | 16 |
| Isolate 19 | *bla*$_{NDM-5}$ | 16 |
| Isolate 20 | *bla*$_{NDM-5}$ | 6 |
| Isolate 21 | *bla*$_{NDM-5}$ | 12 |
| Isolate 22 | *bla*$_{NDM-5}$ | 4 |

**Table S4. Metagenomic comparisons of closely related strains.**

Table S4 is provided as a separate excel file.
